# Multi-Herbal Extracellular Vesicle and Growth Factor Serum for Eyelash Care: A Pilot Randomized, Double Blind, Placebo-Controlled Clinical Study

**DOI:** 10.64898/2026.09.06.26360639

**Authors:** Tsong-Min Chang, Chung-Chin Wu, Huey-Chun Huang, Ji-Ying Lu, Yu-San Chen, Pei-Lun Kao, Wei-Hsuan Tang, Wang-Ju Hsieh, Luke Tzu-Chi Liu, Ivona Percec, Charles Chen, Tsun-Yung Kuo

**Affiliations:** Department of Applied Cosmetology, HungKuang University, Taichung City 433304, Taiwan; Schweitzer Biotech Company, Taipei City 114066, Taiwan; Department of Medical Laboratory Science and Biotechnology, China Medical University, Taichung City 406040, Taiwan; Division of Plastic Surgery, Department of Surgery, University of Pennsylvania, Philadelphia, PA 19122, USA; College of Science and Technology, Temple University, Philadelphia, PA 19122, USA

## Abstract

Beyond their cosmetic significance, eyelashes are critical for retaining ocular moisture and protection against mechanical debris, environmental stressors, and excessive light. While recombinant growth factors support hair growth through activating key follicular signaling pathways, plant-derived EVs modulate local inflammatory and oxidative stress pathways to optimize the follicular microenvironment. Here we conducted a randomized, double-blind, placebo-controlled pilot study to investigate the synergistic effects of a serum containing recombinant Fc-fusion growth factors (rIGF-1, rFGF-7) and extracellular vesicles (EVs) derived from *Zingiber officinale* (ginger) and *Curcuma longa* (turmeric). Thirty healthy female participants were recruited and allocated to Group A (active serum) or Group B (placebo) at a ratio of 1:1. Participants applied the intervention twice daily for 8 weeks. The VISIA® Skin Analysis System was used during clinical visits to objectively measure eyelash density and average length at baseline, Week 4, and Week 8. Group A exhibited a mean density increase of 3.3% at Week 4 and 4.4% at Week 8 relative to baseline. Eyelash length in Group A exhibited significant increases, reaching 8.1% at Week 4 and 11.0% by Week 8. In contrast, Group B showed slight reductions in length (ࢤ1.7% and −3.7% at Weeks 4 and 8, respectively) and a negligible change in density (+0.8% and −1.0% at Week 4 and 8, respectively). Notably, no periocular skin hyperpigmentation, a frequent adverse event associated with pharmacologic agents such as bimatoprost, was observed. These findings demonstrated the potential of combining long-acting recombinant growth factors with plant-derived EVs as an effective, highly tolerable, non-pharmaceutical cosmetic strategy for eyelash enhancement.

## 1. Introduction

Eyelashes are specialized terminal hairs situated along the eyelid margin, representing a functionally and morphologically unique subset of human hair follicles[1,2]. Beyond their undisputed aesthetic appeal, eyelashes perform essential physiological roles in preserving ocular surface integrity[1]. Positioned at the palpebral aperture, they divert airflow away from the cornea, reduce the deposition of airborne particulates, and elicit protective blink reflexes via high tactile sensitivity[1]. Anatomically, eyelash follicles comprise standard follicular components, including the hair shaft, root, bulb, matrix region, and dermal papilla, yet differ markedly from scalp follicles by displaying a substantially shorter anagen (growth) phase and a slower elongation rate[1,2]. Human eyelashes elongate at approximately 0.12-0.14 mm per day, featuring an anagen duration of only 4-10 weeks within a total follicular cycle of 4-11 months[1]. These biological characteristics account for the inherently limited length of eyelashes and highlight the potential utility of topical strategies engineered to extend anagen duration or stimulate follicular activity[1,2].

In addition to their protective capacity, eyelashes are fundamentally linked to perceived facial attractiveness and eye prominence[1]. Longer, denser, and darke eyelashes are widely viewed as accentuating eye expression, driving consumer demand for cosmetic enhancements and growth-directed treatments[3–5]. The therapeutic success of bimatoprost, a prostamide analogue approved for eyelash hypotrichosis, demonstrates that eyelash follicles are biologically responsive mini-organs capable of responding to topical pharmacologic or biologically active signals[3–5]. Mechanistically, bimatoprost promotes lash growth primarily by prolonging the anagen phase and recruitment. However, its clinical utility is frequently limited by adverse effects, most notably undesirable periocular hyperpigmentation and iris color changes [3–5]. There remains a compelling need for targeted, non-pharmaceutical interventions capable of enhancing visible eyelash parameters without triggering localized side effects[3–5].

Insulin-like growth factor-1 (IGF-1) is a growth-promoting peptide involved in hair follicle proliferation, cell survival, and anagen maintenance[6]. In human hair follicle organ culture, IGF-1 stimulates hair shaft elongation at physiological concentrations while delaying catagen transition[7,8]. IGF-1 has also been shown to help maintain anagen characteristics and delay catagen-like changes in cultured follicles[7–9]. Mechanistically, IGF-1 drives anti-apoptotic responses and follicular keratinocyte proliferation via activation of the PI3K/Akt and MAPK/ERK signaling cascades[6,9,10]. Although direct clinical studies assessing IGF-1 in eyelash growth remain sparse, the structural similarities between scalp and eyelash follicles provide a compelling rationale for evaluating topical IGF-1 as a supportive agent for eyelash growth[2,6–10].

Fibroblast growth factor-7 (FGF-7), also known as keratinocyte growth factor (KGF), plays an essential role in epithelial maintenance and follicular morphogenesis[11–13]. FGF-7 is mainly produced by mesenchymal cells and acts on epithelial cells through FGFR2b-mediated signaling[11]. In hair follicles, this mesenchymal-to-epithelial signaling occurs between dermal papilla cells (mesenchymal) and follicular keratinocytes (epithelial), helping regulate follicle development, cycling, and hair shaft production[11–13]. Preclinical models indicate that endogenous KGF/FGF-7 is required for normal follicle differentiation[12,13]. Given that eyelash follicles depend heavily on dermal papilla–epithelial crosstalk, topical FGF-7 may foster a favorable microenvironment for sustained keratinocyte proliferation and eyelash elongation[11–13].

Plant-derived exosome-like nanoparticles (extracellular vesicles, EVs) have emerged as biocompatible nanoscale carriers containing diverse bio-cargo, including lipids, proteins, microRNAs, and secondary metabolites[14–16]. Characterized by low cytotoxicity and favorable cellular uptake, these nano-vesicles participate in intercellular signaling and tissue homeostasis[14]. Ginger(*Zingiber officinale*)-derived EVs represent a prominent class of herb-derived vesicles shown to modulate macrophage-mediated inflammatory responses[15]. They contain bioactive cargo and have been reported to modulate inflammatory responses, including macrophage-related inflammatory pathways[14,15]. A recent PubMed-indexed study specifically investigated ginger-derived extracellular vesicles in an alopecia model, suggesting that ginger vesicle-derived particles may be relevant to hair-loss research[17]. For eyelash enhancement, ginger-derived exosome-like nanoparticles may therefore serve as a supportive botanical nanocomponent by modulating the local follicular microenvironment, particularly inflammation- and stress-related factors that may influence follicle performance[14,15,17].

Similarly, turmeric (*Curcuma longa*)-derived EVs carry bioactive polyphenols, including curcumin, alongside native vesicular lipids and proteins[16,18]. Turmeric-derived nanoparticles have been characterized as vesicle-like structures containing proteins, lipids, nucleic acids, and small molecules, including turmeric-associated bioactive components[16]. Turmeric and its major polyphenol curcumin are widely recognized for anti-inflammatory and antioxidant activities, and turmeric-derived nanoparticles have been reported in experimental models to suppress inflammatory signaling and support tissue repair[18,19]. Although evidence directly linking turmeric-derived exosome-like nanoparticles to eyelash growth is not yet established, their anti-inflammatory and antioxidant properties may provide a rational supportive role in a topical eyelash formulation, especially when combined with follicle-directed growth factors[16,18,19].

Taken together, engineered growth factors (rIGF-1 and rFGF-7) offer targeted direct stimulation of follicular proliferation and anagen signaling, whereas plant-derived EVs (*Z. officinale* and *C. longa*) offer complementary microenvironmental conditioning by mitigating oxidative and inflammatory stress[6–19]. Despite growing interest in growth factor biology and nano-vesicular cosmetics, robust clinical evidence evaluating their combined application on human eyelashes remains limited[2,6–19]. Therefore, this pilot clinical trial was designed to evaluate the clinical efficacy, objective parameters, and local safety profile of an eyelash-enhancing serum incorporating recombinant long-acting growth factors and dual plant-derived EVs.

## 2. Materials and Methods

### 2.1. Participants and Study Design

This prospective, randomized, double-blind, placebo-controlled clinical trial evaluated the efficacy and local tolerability of a multi-herbal EV and growth factor cosmetic serum on eyelashes and periorbital skin. Thirty healthy adult female participants aged 18 to 60 years were enrolled and allocated to either the active serum group (Group A) or placebo group (Group B) at a 1:1 ratio (*n* = 15 per group). Inclusion criteria include healthy female adults aged 18 to 60 years (inclusive), who do not have any chronic diseases, major illnesses, or allergic conditions. Furthermore, eligible participants must not have taken any medication or used any eyelash care products within the past month. Exclusion criteria include currently pregnant or breastfeeding participants, or having any chronic diseases, major illnesses, or allergic conditions. Additionally, individuals are ineligible if they are currently using eyelash care or growth products, are already enrolled in another cosmetic test for eyelash care, or have undergone any medical aesthetic procedures in the eye area within the past three months. The trial also excludes participants with existing eye injuries or active eye diseases, such as conjunctivitis, blepharitis, or hordeolum..

Prior to enrollment, all participants underwent a 24-hour patch test on the inner forearm to screen for topical sensitivity. Participants must thoroughly cleanse their faces and remove contact lenses if worn before application of the intervention. Following their routine skincare regimen, participants should apply a thin layer of an appropriate amount of intervention along the roots of the upper eyelid lashes, taking care to avoid the eyelashes themselves and prevent contact with the eyes. Any excess serum must be wiped away. The product is to be applied twice daily, in the morning and evening. After application, participants should wait a few minutes to allow absorption, during which time they must avoid water contact and rubbing their eyes. Participants are required to self-record the timing of each product use. The duration of intervention was 8 weeks (56 days), with measurements taken at baseline (Week 0/Day 0) and in Weeks 4 (Day 28) and 8 (Day 56).

Descriptions of interventions are as follows:

- Group A (Test product): Base formula containing *Zingiber officinale* root-derived extracellular vesicles (INCI ID: 38118) ,*Curcuma longa* rhizome vesicles (INCI ID: 40153), long-acting recombinant human IGF-1 (rIGF-1; INCI ID: 40916), and long-acting recombinant human FGF-7 (rFGF-7; INCI ID: 40201).

- Group B (Placebo Control): Identical vehicle base lacking active recombinant growth factors and plant EVs.

*Z. officinale* and *C. longa* extracellular vesicles were isolated using a previously described centrifugation and filtration method[20]. rIGF-1 (INCI ID: 40916) and rFGF-7 (INCI ID: 40201) are recombinant proteins modified for long-acting by fusing human IGF-1 or human FGF-7 to a human IgG1 Fc region fragment with a flexible linker described previously[21].

### 2.2. Randomization and Blinding

Participants were assigned (1:1 ratio; *n* = 15/group) using block randomization with a fixed block size of two. An independent coordinator not involved in participant recruitment or clinical evaluation prepared sequentially numbered, opaque, tamper-evident sealed envelopes using a random allocation sequence generated by the principal investigator. Envelopes were stored securely and opened only after written informed consent and baseline eligibility were secured.

Double-blind integrity was maintained throughout the trial: participants, trial coordinators, clinical assessors, and biostatisticians remained masked to group assignments until database lock. The test serum and placebo were provided in identical primary packaging, matching in color, odor, viscosity, texture, and labeling. Units were designated solely by participant identification numbers. Emergency unblinding procedures were established but not required.

### 2.3. Assessment of Outcomes

Primary outcomes were assessed with a non-invasive diagnostic system from three angles of the face (left, right, and front) using the VISIA® Skin Analysis System (Canfield Imaging Systems, Fairfield, NJ, USA), which uses objective computer vision analysis with standardized, high-resolution imaging (IntelliFlash® white-light and head positioning).

• Eyelash Density (Pixel Area):: The system uses an image segmentation algorithm to isolate dark eyelash pixels from the lighter skin background. It calculates the total pixel area occupied by eyelashes within a specified Region of Interest (ROI) to determine density, expressed as total eyelash pixels/ROI.

• Average Eyelash Length (mm): Automated feature recognition traced individual lashes from follicular origin to distal tip. Digital pixel distances were calibrated and converted to physical length (mm).

• The results were reported as a percentage change from the baseline value, as in the following formula:

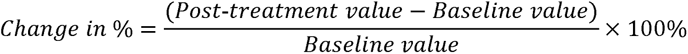

The results were calculated as the mean of measurements from the left, right, and front regions for each participant, then converted to a percentage change from baseline.

### 2.4. Statistical Analysis

Statistical analyses were performed using the built-in statistical package of GraphPad Prism 6.01 (Boston, MA, USA). Results were presented as mean values with standard error of the mean (SEM). For comparing demographic baseline values, the χ^2^ test was used for gender and the unpaired t-test was used for continuous variables. To compare within-group changes over time, a paired t-test was used. For between-group comparisons at each time point, an unpaired t-test was used. For age-stratified subgroup analysis (cut-off as > 40 and ≤ 40 years), an unpaired t-test was used to calculate the significance between the groups, while Pearson correlation *r* was used to calculate the correlation between participants’ age and change in length/density. A *p*-value < 0.05 was considered statistically significant. All tests were two-tailed. The sample size in this clinical study was not based on any statistical assumptions or calculations, as this was an exploratory study aimed at observing the effects of the formulations.

## 3. Results

Between 1 November 2025 and 26 May 2026, 30 eligible female participants were enrolled and all were screened for negative for irritation or allergic reactions to the test product by patch test prior to the study. The CONSORT Flow Diagram of the study participants is shown in Figure 1. The trial recorded zero attrition, with no subjects lost to follow-up or discontinued during the eight-week period (100% retention rate), which demonstrated the tolerability of the intervention. Table 1 summarizes the demographic characteristics of the participants, including age, eyelash length, and density at the start of the study, which showed no significant differences in baseline characteristics between the two groups.

**Figure 1.**
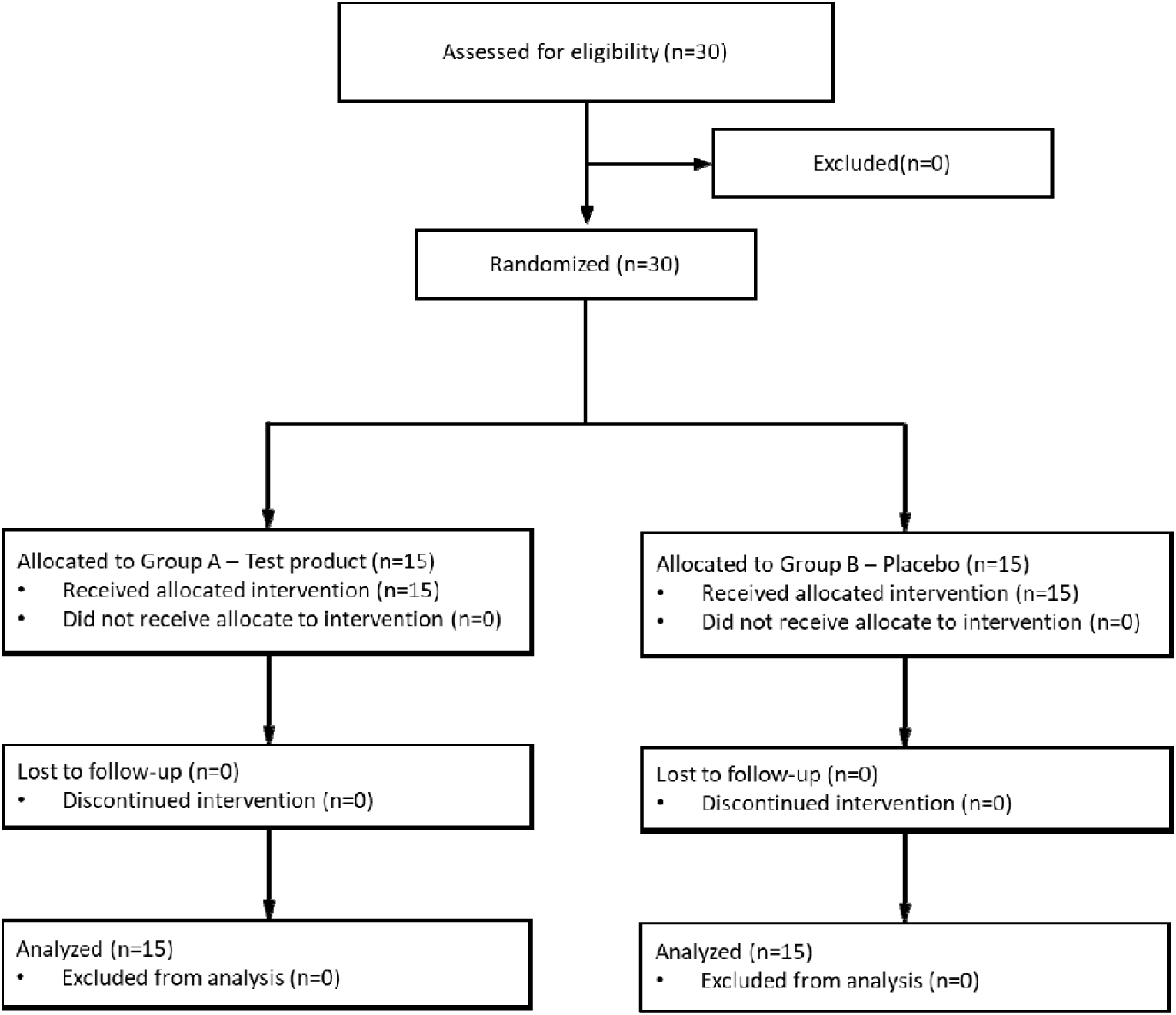
**CONSORT flowchart of the study.**

**Table 1.**
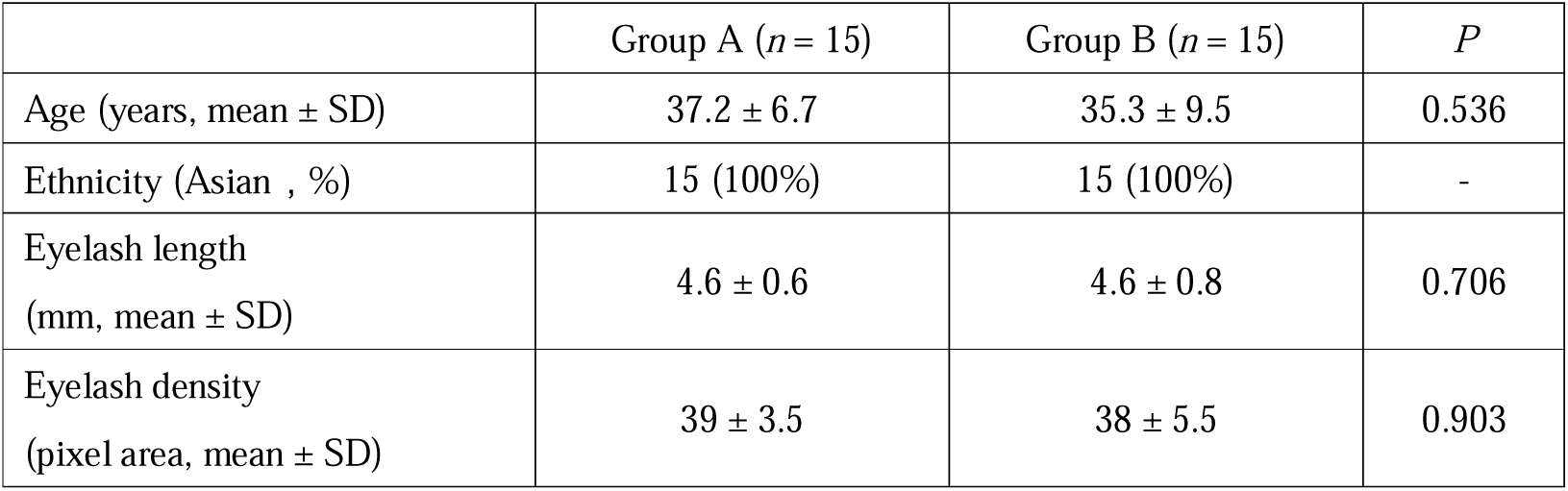
Demographic characteristics of the participants in each of the groups. Statistical significance (*P*) between the two groups was calculated with a two-sample t-test.

The participants were assessed for changes in eyelash length and density as primary outcomes of the study. In Group A (active serum), eyelash density showed a progressive upward trajectory, significantly increasing (*p* < 0.0001) to 3.27% and 4.43% compared to baseline at Week 4 and 8, respectively (Figures 2 and 3). In contrast, Group B (placebo) had minimal change in density at Week 4 (0.76%) and even decreased at Week 8 (ࢤ0.95%) when compared to baseline (Figures 2 and 3). Group A had significantly (*p* < 0.0001) longer eyelashes at Week 4 and 8 by 8.07% and 10.99% compared to baseline length (Figures 2 and 3). Placebo did not have any positive effects on eyelash length; in fact, the length decreased to −1.70% at Week 4 and −3.71% at Week 8 compared to baseline, although neither change was statistically significant (Figures 2 and 3). These findings also revealed a significant disparity in early performance. Even at the four-week midpoint, the eyelashes lengthened in the serum group (+8.07%) whereas eyelashes shortened in the placebo group (ࢤ1.70%). This indicates that the active formulation provides an accelerated response in follicular density, differentiating the test product’s efficacy from the natural shortening of the eyelashes and placebo effects. Representative photographs taken with the VISIA® System during clinical visits at baseline (Week 0) and end of study (Week 8) are shown in Figure 4. Interestingly, no hyperpigmentation in the periocular region is seen after 8 weeks of twice-daily serum use.

**Figure 2.**
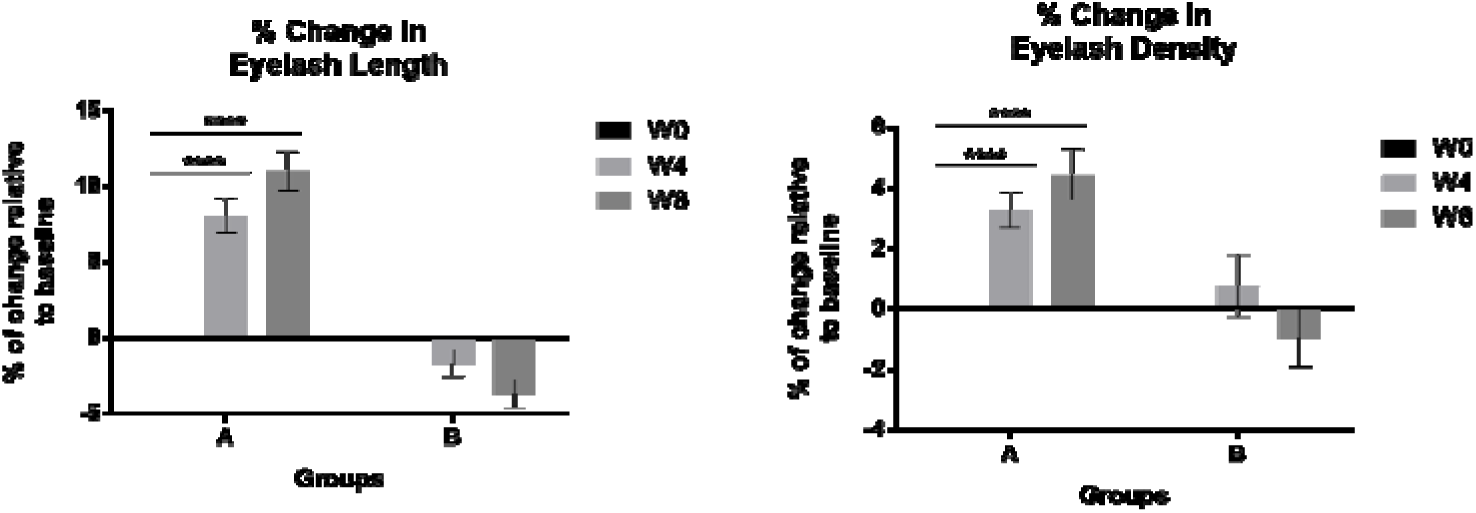
Change in mean eyelash length (left) and density (right) by group at 0, 4, and 8 weeks after intervention. Statistical significance between the two time points within the group was calculated with paired t-test. **** *p* < 0.0001.

**Figure 3.**
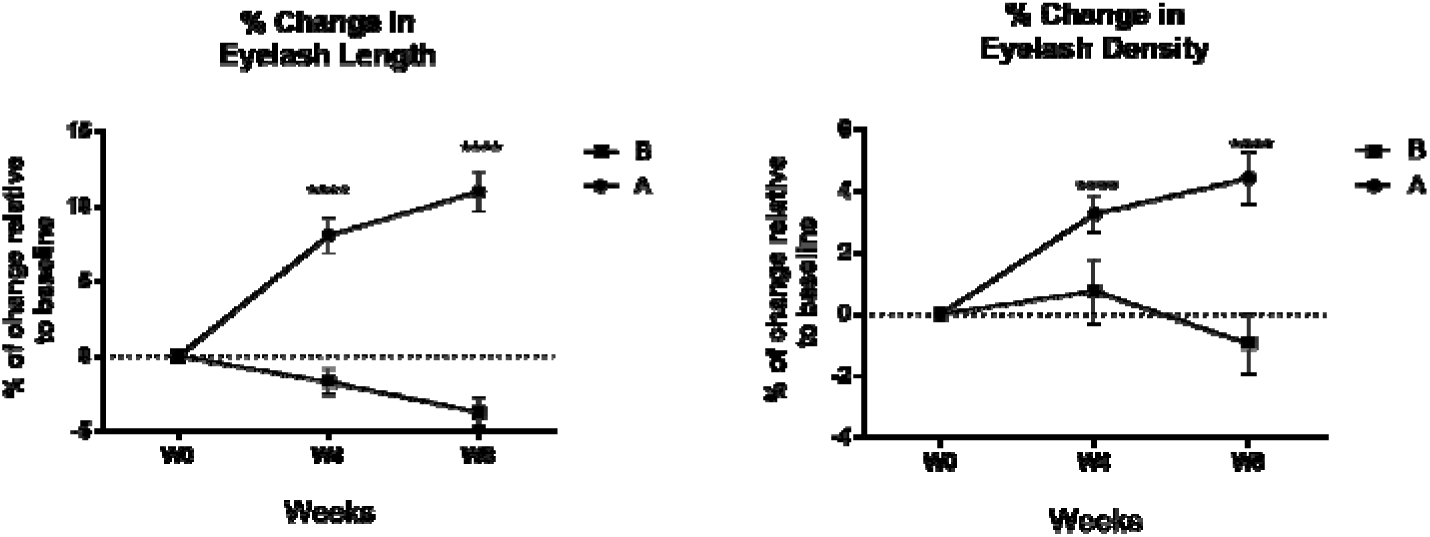
Change in mean eyelash length (left) and density (right) by group at 0, 4, and 8 weeks after intervention. Statistical significance between the two groups at the same time point was calculated with a two-sample t-test. **** *p* < 0.0001.

**Figure 4.**
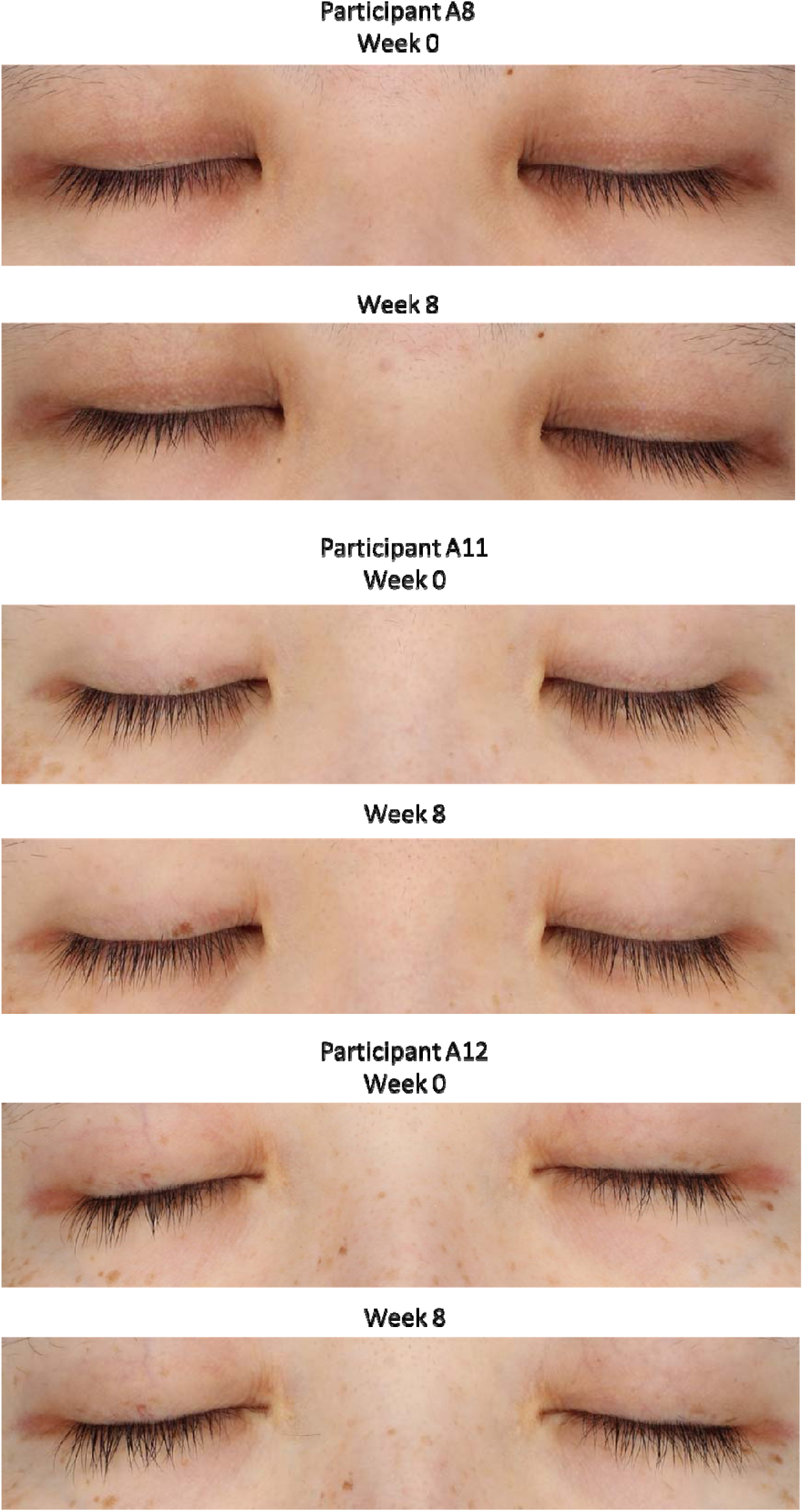
Representative images of four participants from Group A (serum) at Weeks 0 and 8 of the study captured by the VISIA® system.

Although the present study sample size is underpowered for subgroup analyses, we nevertheless performed exploratory subgroup analysis to assess whether age (cutoff of 40 years) could have an apparent effect on the treatment response in the treatment group. For eyelash length, no significant difference in percent change was observed between participants > 40 years (*n* = 6) and ≤ 40 years (*n* = 9) at either Week 4 (9.47% vs. 7.14%, *p* = 0.34) or Week 8 (12.17% vs. 10.21%, *p* = 0.48), and age showed no correlation with treatment response when analyzed continuously across the full cohort (*r* ≈ −0.01 to −0.04, *p* > 0.90). For eyelash density, participants > 40 years unexpectedly showed a numerically larger increase than their younger counterparts at Week 4 (4.68% vs. 2.33%, *p* = 0.048) and Week 8 (6.05% vs. 3.35%, *p* = 0.12), and the correlation between age and density was borderline significant and had a positive value (*r* = 0.51, *p* = 0.051). A similar age-associated pattern observed in the placebo group was not considered a genuine age effect, as it was heavily influenced by a single outlier, given the small sample size (*n* = 4 for > 40 years) and by a significant inverse relationship between baseline density and percent change (*r* = −0.62, *p* = 0.01), which could be due to regression toward the mean, rather than a true biological trend.

## 4. Discussion

To our knowledge, this study provides the first randomized, controlled clinical study to evaluate the combined efficacy of engineered, long-acting recombinant growth factors and plant-derived EVs specifically optimized for eyelashes. IGF-1 is essential for maintaining anagen characteristics within the eyelash follicle by activating the PI3K/Akt and MAPK/ERK pathways, which are the primary drivers of cell proliferation and survival [6, 9]. By sustaining follicular viability and prolonging the anagen window, rIGF-1 delays the transition to catagen-like changes, effectively keeping a higher percentage of follicles in the active growth phase for longer periods [7, 8]. FGF-7 complements the action of IGF-1 by facilitating critical mesenchymal-to-epithelial signaling. Simultaneously, FGF-7 acts via epithelial FGFR2b receptor signaling, stimulating dermal papilla-keratinocyte crosstalk essential for epithelial cell proliferation and hair shaft elongation [11]. This interaction is a primary regulator of hair shaft production and follicle development [12, 13]. By reinforcing the proliferative capacity of the epithelial matrix, rFGF-7 supports the increase in hair shaft formation observed in our trial.

The targeted growth factor signaling employed in this study was recombinant growth factor proteins specifically engineered for prolonged bioactivity. Growth factors have a short half-life of typically minutes and are prone to degradation by the complex skin and external environment, which limits their usefulness in topical use [22,23]. Unlike the short half-life of native growth factors, the rIGF-1 and rFGF-7 used in this serum are fused to a human IgG1 Fc fragment to enhance stability and duration, thereby addressing the short-lived nature of typical topical proteins[24,25]. While standard mechanistic models note the theoretical relationship between cell growth and cellular transformation, extensive animal and clinical data consistently demonstrate that topical application is safe and does not induce oncogenic transformation or systemic toxicity [26–28]. This favorable safety profile is largely attributed to the localized, transient nature of topical growth factors, even with the long-acting recombinant Fc-fusion proteins engineered for this study. [6]. To ensure the highest standards of clinical responsibility and proactive mitigation, individuals with a history of skin malignancies may benefit from consultation with physicians prior to use, and ongoing long-term evaluation will continue to reinforce the established safety profile of these novel recombinant growth factors.

To complement growth factor signaling, the formula incorporates plant-derived EVs from ginger (Z. officinale) and turmeric (C. longa). These nanovesicles provide natural lipid bilayer protection for sensitive microRNAs, proteins, and polyphenols, facilitating efficient delivery to the follicular microenvironment [14, 16]. Ginger-derived EVs are known to contribute to follicle stability by modulating the local environment and macrophage-related inflammatory pathways, mitigating micro-inflammation and stress-related factors that can lead to premature follicular regression [14, 15, 17]. Concurrently, turmeric-derived EVs provide essential antioxidant and anti-inflammatory support for a favorable microenvironment to promote hair growth [18, 19]. The inclusion of curcumin-associated bioactive components within these vesicles protects delicate follicular structures from oxidative damage, which is vital for maintaining the health of both the periorbital skin and the melanocytes responsible for eyelash pigment. While this study compared the combination of EVs and growth factors against a placebo lacking all active ingredients, it does not permit attribution of the observed effect to any individual component or to their interaction. It remains possible that the recombinant growth factors alone, the plant-derived EVs alone, or a synergistic interaction between the two ingredient classes are responsible for the clinical response observed.

Currently, bimatoprost remains the only FDA-approved pharmaceutical agent for eyelash hypotrichosis [3, 4]. While effective at extending the anagen phase, its long-term clinical use is limited by side effects such as periorbital and iris hyperpigmentation [5]. Hyperpigmentation by prostaglandin analogs is attributed to prostaglandin F_2_α receptor-mediated stimulation of melanogenesis, including upregulation of tyrosinase activity and increased melanocyte dendricity, rather than promoting true melanocyte proliferation[29,30]. Furthermore, iris pigmentation changes associated with prostamides are often permanent [31]. In contrast, the present formulation contains no prostaglandin analogs and instead offers a non-pharmaceutical, multi-pathway approach using recombinant growth factors and plant-EVs to mediate follicular signaling. This mechanism also distinguishes the formulation within the expanding over-the-counter (OTC) cosmetic market. While conventional OTC serums typically rely on simple peptides or botanicals for lash conditioning, others covertly incorporate synthetic prostamide/prostaglandin analogs despite “natural” market positioning[32,33]. Furthermore, unlike FDA-regulated pharmaceuticals subject to rigorous New Drug Application reviews, OTC cosmetics face no pre-market FDA safety or efficacy validation and manufacturers must be responsible for the safety and efficacy of the products, at the same time without claiming therapeutic benefits[34]. Thus, the present formulation occupies a distinct niche in the current eyelash treatment landscape: it contains no prostaglandin-class actives but instead utilizes long-acting recombinant growth factors (rIGF-1, rFGF-7) engineered to directly engage anagen-promoting signaling pathways, and supported by plant-derived EVs that modulate the follicular microenvironment. This mechanistically targeted action uniquely positions the serum as an alternative that is neither prescription pharmaceutical nor conventional OTC cosmetic/prostaglandin treatment.

An interesting observation in our study was the mild decline in lash length and density observed in the placebo cohort over the 8-week period. Trial enrollment spanned November through May, capturing a seasonal shift from late autumn to spring. Human scalp hair displays well-documented seasonal shedding cycles, characterized by peak anagen ratios in spring and increased telogen shedding during late summer/autumn [35]. Although seasonal dynamics in human eyelashes are less characterized, eyelash follicles share fundamental biological properties with scalp follicles [1,2]. It may remain a possibility that enrollment timing contributed to individual variability in growth response and may partly explain the finding that the placebo group exhibited a decrease in eyelash length and density rather than remaining unchanged over the study period. This decline could reflect natural cyclical variation in eyelash growth captured incidentally by the study’s timing, which, given a longer study duration, could be fully pictured as an eyelash follicular cycle that typically ranges from 4-11 months[1].

As with exploratory nature of the study, we ought to point out the following limitations. The small sample size (*n* = 15 per group) consisting of exclusively Asian females and the short 8-week duration, while sufficient for an exploratory analysis, limited the generalizability of the findings to a broader population, even though the effects were apparent as early as 4 weeks. Eyelash morphology and growth characteristics have been shown to differ meaningfully by ethnic background, as a study found that Asian eyelashes have a lower curl-up and lift-up angle, fewer lashes per eyelid, and a thicker average lash diameter compared with Caucasian eyelashes[36]. Given these structural differences, the magnitude and time-course of response to a topical growth factor and EV-based serum observed in this Asian female cohort may not be applicable to populations with differing eyelash morphology, and future studies in more ethnically diverse and sex-balanced cohorts are warranted to establish the generalizability of this formulation’s efficacy. Furthermore, while the current protocol focused on an 8-week endpoint to align with the eyelash anagen phase, the complexity of the full 4- to 11-month follicular cycle warrants extended observation, as well as seasonal variation noted above [1, 2]. In addition, head-to-head comparisons against bimatoprost or other prostamide analogues would further define the serum’s position in the market. As mentioned previously, the safety of long-term use of growth factors would also require painstaking evaluation and monitoring to avoid malignant transformation.

The results of this clinical trial affirmed that the synergistic actions of long-acting rIGF-1, rFGF-7, and plant-derived EVs from ginger and turmeric represent a high-value, effective, and well-tolerated approach to eyelash enhancement. The objective data from the VISIA® system, paired with the absence of ocular irritation, position this formulation as a cosmetic alternative to traditional pharmacological treatments for eyelash hypotrichosis.

## Author Contributions

Conceptualization, supervision and project administration: T.-M.C., C.C., T.-Y.K. Visualization and formal analysis: T.-M.C., W.-J.H., L.T.-C.L. Investigation, data acquisition and curation: T.-M.C., H.-C.H., J.-Y.L. Methodology: T.-M.C., C.-C.W., H.-C.H., J.-Y.L., Y.-S.C., P.-L.K., W.-H.T., I.P., T.-Y.K. Resources: C.-C.W., Y.-S.C., P.-L.K., W.-H.T., T.-Y.K. Writing—original draft preparation: T.-M.C., W.-J.H., L.T.-C.L. Writing—review and editing: T.-M.C., W.-J.H., L.T.-C.L., I.P., C.C., T.-Y.K. All authors reviewed the manuscript.

## Funding

Schweitzer Biotech Company provided the necessary funding for the study as well as provided the test products, but had no role in enrollment, participation, data collection, data analysis, or completion of final report of the clinical study. All participants provided written informed consent prior to the start of the study.

## Institutional Review Board Statement

The finalized protocol, case report form, advertisement, and informed consent form were reviewed and approved by the institutional review board of the Antai Tian-Sheng Memorial Hospital (IRB No. 25-089-A, approved 21 October, 2025). All participants provided written informed consent prior to the start of the study. This study was conducted in compliance with the Declaration of Helsinki and in compliance with all International Conference on Harmonization Good Clinical Practice Guidelines. This clinical study was registered on ClinicalTrials.gov under NCT07271199 on 27 November, 2025.

## Data Availability Statement

All relevant data is provided within the manuscript.

## Data Availability

All relevant data is provided within the manuscript.

## Acknowledgments

We thank colleagues at Schweitzer Biotech Company for reviewing and providing feedback for the manuscript preparation.

## Conflicts of Interest

Authors C.-C. W., Y.-S. C., P.-L. K., W.-H. T., W.-J. H., L. T.-C. L., C. C., and T.-Y. K. are employed by Schweitzer Biotech Company. T.-M. C. and I. P. are consultants employed by Schweitzer Biotech Company. The remaining authors declare that the research was conducted in the absence of any commercial or financial relationships that could be construed as a potential conflict of interest.

## Notes

### Clinical Trial

NCT07271199

### Author Declarations

The finalized protocol, case report form, advertisement, and informed consent form were reviewed and approved by the institutional review board of the Antai Tian-Sheng Memorial Hospital (IRB No. 25-089-A, approved 21 October, 2025). All participants provided written informed consent prior to the start of the study. This study was conducted in compliance with the Declaration of Helsinki and in compliance with all International Conference on Harmonization Good Clinical Practice Guidelines.

## References

1. Aumond, S.; Bitton, E. The eyelash follicle features and anomalies: A review. J Optom 2018, 11, 211–222, doi:10.1016/j.optom.2018.05.003.

2. Paus, R.; Burgoa, I.; Platt, C.I.; Griffiths, T.; Poblet, E.; Izeta, A. Biology of the eyelash hair follicle: an enigma in plain sight. Br J Dermatol 2016, 174, 741–752, doi:10.1111/bjd.14217.

3. Cohen, J.L. Enhancing the growth of natural eyelashes: the mechanism of bimatoprost-induced eyelash growth. Dermatol Surg 2010, 36, 1361–1371, doi:10.1111/j.1524-4725.2010.01522.x.

4. Law, S.K. Bimatoprost in the treatment of eyelash hypotrichosis. Clin Ophthalmol 2010, 4, 349–358, doi:10.2147/opth.s6480.

5. Fagien, S. Management of hypotrichosis of the eyelashes: Focus on bimatoprost. Clin Cosmet Investig Dermatol 2010, 3, 39–48, doi:10.2147/ccid.s5488.

6. Hsieh, W.J.; Qiu, W.Y.; Percec, I.; Chang, T.M. Insulin-like Growth Factor 1 (IGF-1) in Hair Regeneration: Mechanistic Pathways and Therapeutic Potential. Curr Issues Mol Biol 2025, 47, doi:10.3390/cimb47090773.

7. Philpott, M.P.; Sanders, D.A.; Kealey, T. Effects of insulin and insulin-like growth factors on cultured human hair follicles: IGF-I at physiologic concentrations is an important regulator of hair follicle growth in vitro. J Invest Dermatol 1994, 102, 857–861, doi:10.1111/1523-1747.ep12382494.

8. Philpott, M.P.; Sanders, D.; Westgate, G.E.; Kealey, T. Human hair growth in vitro: a model for the study of hair follicle biology. J Dermatol Sci 1994, 7 *Suppl*, S55–72, doi:10.1016/0923-1811(94)90036-1.

9. Ahn, S.Y.; Pi, L.Q.; Hwang, S.T.; Lee, W.S. Effect of IGF-I on Hair Growth Is Related to the Anti-Apoptotic Effect of IGF-I and Up-Regulation of PDGF-A and PDGF-B. Ann Dermatol 2012, 24, 26–31, doi:10.5021/ad.2012.24.1.26.

10. Bassino, E.; Gasparri, F.; Giannini, V.; Munaron, L. Paracrine crosstalk between human hair follicle dermal papilla cells and microvascular endothelial cells. Exp Dermatol 2015, 24, 388–390, doi:10.1111/exd.12670.

11. Zheng, Y.; Liu, W.H.; Yang, B.; Milman Krentsis, I. Primer on fibroblast growth factor 7 (FGF 7). Differentiation 2024, 139, 100801, doi:10.1016/j.diff.2024.100801.

12. Danilenko, D.M.; Ring, B.D.; Yanagihara, D.; Benson, W.; Wiemann, B.; Starnes, C.O.; Pierce, G.F. Keratinocyte growth factor is an important endogenous mediator of hair follicle growth, development, and differentiation. Normalization of the nu/nu follicular differentiation defect and amelioration of chemotherapy-induced alopecia. Am J Pathol 1995, 147, 145–154.

13. Guo, L.; Degenstein, L.; Fuchs, E. Keratinocyte growth factor is required for hair development but not for wound healing. Genes Dev 1996, 10, 165–175, doi:10.1101/gad.10.2.165.

14. Kim, J.; Li, S.; Zhang, S.; Wang, J. Plant-derived exosome-like nanoparticles and their therapeutic activities. Asian J Pharm Sci 2022, 17, 53–69, doi:10.1016/j.ajps.2021.05.006.

15. Zhu, H.; He, W. Ginger: a representative material of herb-derived exosome-like nanoparticles. Front Nutr 2023, 10, 1223349, doi:10.3389/fnut.2023.1223349.

16. Wei, Y.; Cai, X.; Wu, Q.; Liao, H.; Liang, S.; Fu, H.; Xiang, Q.; Zhang, S. Extraction, Isolation, and Component Analysis of Turmeric-Derived Exosome-like Nanoparticles. Bioengineering (Basel) 2023, 10, doi:10.3390/bioengineering10101199.

17. Hao, Y.; Yang, Q.; Zhang, H.; Bai, C.; Liu, X.; Gao, Y. Ginger-Derived Extracellular Vesicles: A Natural Solution for Alopecia. Curr Drug Deliv 2026, 23, 111–124, doi:10.2174/0115672018321133240829074400.

18. Liu, C.; Yan, X.; Zhang, Y.; Yang, M.; Ma, Y.; Zhang, Y.; Xu, Q.; Tu, K.; Zhang, M. Oral administration of turmeric-derived exosome-like nanovesicles with anti-inflammatory and pro-resolving bioactions for murine colitis therapy. J Nanobiotechnology 2022, 20, 206, doi:10.1186/s12951-022-01421-w.

19. Hewlings, S.J.; Kalman, D.S. Curcumin: A Review of Its Effects on Human Health. Foods 2017, 6, doi:10.3390/foods6100092.

20. Chang, T.M.; Wu, C.C.; Huang, H.C.; Lu, J.Y.; Chuang, C.H.; Kao, P.L.; Tang, W.H.; Liu, L.T.C.; Qiu, W.Y.; Percec, I. A 28-Day Pilot Study of the Effects on Facial Skin Hydration, Elasticity, and Texture of a Centella asiatica Extracellular Vesicle-Based Skin Care Formulation. Cosmetics 2025, 12, 186.

21. Chang, T.M.; Wu, C.C.; Huang, H.C.; Lu, J.Y.; Chuang, C.H.; Kao, P.L.; Tang, W.H.; Hsieh, W.J.; Liu, L.T.C.; Qiu, W.Y. Centella asiatica L. Urb. Extracellular Vesicle and Growth Factor Essence for Hair and Scalp Health: A 56-Day Exploratory Randomized Trial. Cosmetics 2025, 12, 253.

22. Senekowitsch-Schmidtke, R.; Steiner, K.; Haunschild, J.; Mollenstadt, S.; Truckenbrodt, R. In vivo evaluation of epidermal growth factor (EGF) receptor density on human tumor xenografts using radiolabeled EGF and anti-(EGF receptor) mAb 425. Cancer Immunol Immunother 1996, 42, 108–114, doi:10.1007/s002620050259.

23. Rajpathak, S.N.; Gunter, M.J.; Wylie-Rosett, J.; Ho, G.Y.; Kaplan, R.C.; Muzumdar, R.; Rohan, T.E.; Strickler, H.D. The role of insulin-like growth factor-I and its binding proteins in glucose homeostasis and type 2 diabetes. Diabetes Metab Res Rev 2009, 25, 3–12, doi:10.1002/dmrr.919.

24. Zhang, W.; Zhang, D.; Shen, M.; Liu, Y.; Tian, Y.; Thomson, A.W.; Lee, W.P.; Zheng, X.X. Combined administration of a mutant TGF-beta1/Fc and rapamycin promotes induction of regulatory T cells and islet allograft tolerance. J Immunol 2010, 185, 4750–4759, doi:10.4049/jimmunol.1000769.

25. Mohammadi, A.; Heydari, M.A.; Jamalpoor, Z. Growth factors and cytokines for regenerative medicine: translational challenges and emerging engineering solutions. Regen Eng Transl Med 2025, 1–22.

26. Baserga, R.; Sell, C.; Porcu, P.; Rubini, M. The role of the IGF-I receptor in the growth and transformation of mammalian cells. Cell Prolif 1994, 27, 63–71, doi:10.1111/j.1365-2184.1994.tb01406.x.

27. Ropiquet, F.; Huguenin, S.; Villette, J.M.; Ronfle, V.; Le Brun, G.; Maitland, N.J.; Cussenot, O.; Fiet, J.; Berthon, P. FGF7/KGF triggers cell transformation and invasion on immortalised human prostatic epithelial PNT1A cells. Int J Cancer 1999, 82, 237–243, doi:10.1002/(sici)1097-0215(19990719)82:2<237::aid-ijc14>3.0.co;2-q.

28. Huang, H.C.; Hsieh, W.J.; Percec, I.; Chang, T.M. Fibroblast Growth Factor-7 and Hair Biology: Bridging Basic Science and Therapeutic Applications. Curr Issues Mol Biol 2026, 48, doi:10.3390/cimb48010102.

29. Lee, H.E.; Lim, S.K.; Im, M.; Kim, C.D.; Seo, Y.J.; Lee, J.H.; Lee, Y. Hypertrichosis and Hyperpigmentation in the Periocular Area Associated with Travoprost Treatment. Ann Dermatol 2015, 27, 637–638, doi:10.5021/ad.2015.27.5.637.

30. Wand, M.; Ritch, R.; Isbey, E.K., Jr.; Zimmerman, T.J. Latanoprost and periocular skin color changes. Arch Ophthalmol 2001, 119, 614–615.

31. Stjernschantz, J.W.; Albert, D.M.; Hu, D.N.; Drago, F.; Wistrand, P.J. Mechanism and clinical significance of prostaglandin-induced iris pigmentation. Surv Ophthalmol 2002, 47 *Suppl 1*, S162–175, doi:10.1016/s0039-6257(02)00292-8.

32. Baiyasi, M.; St Claire, K.; Hengy, M.; Tur, K.; Fahs, F.; Potts, G. Eyelash serums: A comprehensive review. J Cosmet Dermatol 2024, 23, 2328–2344, doi:10.1111/jocd.16278.

33. RevitaLash Advanced Eyelash Conditioner - ingredients. Available online: https://incidecoder.com/products/revitalash-advanced-eyelash-conditioner (accessed on 14 July 2026).

34. DRUG, U.S.F. Cosmetics & U.S. Law. Available online: https://www.fda.gov/cosmetics/cosmetics-laws-regulations/cosmetics-us-law (accessed on 24 July 2026).

35. Randall, V.A.; Ebling, F.J. Seasonal changes in human hair growth. Br J Dermatol 1991, 124, 146–151, doi:10.1111/j.1365-2133.1991.tb00423.x.

36. Na, J.I.; Kwon, O.S.; Kim, B.J.; Park, W.S.; Oh, J.K.; Kim, K.H.; Cho, K.H.; Eun, H.C. Ethnic characteristics of eyelashes: a comparative analysis in Asian and Caucasian females. Br J Dermatol 2006, 155, 1170–1176, doi:10.1111/j.1365-2133.2006.07495.x.

